# A policy for delivery of essential medicines to vulnerable population in Argentina: a case study of the REMEDIAR program

**DOI:** 10.64898/2026.06.05.26354987

**Authors:** Maisa Havela, Lucía Bartolomeu, Gisela Bardi, Adolfo Rubinstein

## Abstract

Essential medicines are one of the cornerstones of financial protection and health equity. The REMEDIAR Program is an initiative of the Argentine Ministry of Health aimed at ensuring free access to essential medicines for the uninsured at the point of care in primary healthcare centers (PHC). This study analyzes the financing, procurement, and distribution of this program over two decades (2002–2024). It evaluates how the program’s capacity to navigate economic and political challenges ensured an uninterrupted supply of essential drugs at the primary healthcare level in a federal country where health services are devolved to provinces.

We adopted a mixed-methods approach to examine the duality between international concessional loans and domestic treasury funding. Findings reveal that while international financing enhanced predictability and efficiency—reducing procurement timelines from 458 to 235 days—it also constrained domestic planning through external conditionalities. Conversely, while national centralized procurement achieved superior price efficiency and lower dispersion, it faced rigidities in adapting to local needs.

Territorial distribution analysis confirms that REMEDIAR reduced access barriers for vulnerable households without formal insurance. However, the program entered a stabilization phase, failing to consolidate robust coordination with subnational policies, becoming entrenched in its own operational logic.

The study concludes that program effectiveness depends not only on resource volume but on management quality. To guarantee long-term sustainability, transition to national financing requires profound institutional redesign. This must integrate operational capacities with federal coordination and domestic regulations, ensuring that the primary healthcare supply chain remains resilient to macroeconomic volatility and political shifts, aligned with sub-national strategies.

## Introduction

Argentina is an upper-middle-income federal country where NCDs account for more than 70% of the burden of disease. It has a fragmented and segmented health system divided into three large sectors—the public (35%), social security (55%), and private (10%) —as found in many Latin American countries. The public sector, funded by taxes, has a devolution-type decentralization to the provinces, giving the federal Ministry of Health (MoH) a rather narrow (but strategic) role in national health policy stewardship. Public health funds usually flow from national to provincial to local budgets with no strings attached, leaving the central MoH with little leverage to improve efficiency or accountability or even influence provincial health spending. As a result, the federal level accounts for only 20% of the public sector health expenditures. Although compared with other countries in the region, its health care system performs well on several key indicators, the major health care problems in Argentina today are related to both equity and efficiency, as in many other Latin American countries (1).

The REMEDIAR Program is an initiative of the Argentine Ministry of Health aimed at ensuring free access to essential medicines for the uninsured at the point of care in primary healthcare centers (PHC). It was established in 2002 as part of a strategy to address the healthcare needs of the most vulnerable populations, particularly during Argentina’s dramatic economic and social crisis at the time. Over the years, REMEDIAR has evolved as a key component of Argentinás public health system. The program’s main objectives are: provide medicines by delivering essential ambulatory drugs free of charge for the treatment of common illnesses, primarily at primary health care centers (PHC); strengthen the primary healthcare system by enhancing the capacity of PHC to provide comprehensive and high-quality care; and reduce inequities by ensuring access to medicines in underserved regions and mostly for poorer populations relying only on public coverage. According to the latest census carried out in 2022, more than 16 million people have access only to the public care system (35.8%), and thus are the primary beneficiaries of REMEDIAR program (2).

Throughout its more than two decades of implementation, the program shifted from an emergency strategy to address the 2001–2002 economic crisis to a structural policy within Argentina’s healthcare system. The medicines are distributed in kits containing a defined package of essential drugs to address prevalent conditions such as hypertension, diabetes, asthma, and other chronic conditions, as well as antibiotics for common infections, among other ambulatory drugs. Each PHC receives these kits adjusted for local needs and the population they serve, dispensing the medications to individuals with exclusive public coverage (without social security or private insurance). Over time, changes have been made to its funding sources, procurement modalities, list of essential medicines included in the kits, and prioritized diseases, among other critical aspects. Despite REMEDIAR being a federal program, public health in Argentina is mostly decentralized to the provinces, and in some provinces, to municipalities as well. In this regard, there are significant complementarities with medication coverage policies implemented at the provincial and municipal levels. Historically, the REMEDIAR Program has faced persistent challenges stemming from high inflation, recurrent economic crises, and shifting political priorities. A pivotal juncture in its evolution was the transition from international loan financing to domestic treasury funding. Consequently, REMEDIAR has operated over time under two distinct paradigms: an International Loan-Based Model and a National Program Model. The former, supported by the Inter-American Development Bank (IDB), prioritized structured planning, rigorous monitoring, and international tenders with supplier payments in U.S. dollars. These operations typically followed a *pari passu* cost-sharing logic, in which the IDB financed a substantial portion of the project (typically 50–80%) while the national government provided the remaining local counterpart. In contrast, the National Program Model relies on domestic budgetary allocations, subjecting procurement and financial management to national regulatory frameworks—such as local bids and inter-administrative agreements—with payments executed in local currency and tied to the country’s fiscal space. These models differ significantly in budgetary predictability, procurement efficiency, indicator tracking, and operational adaptability.

The conceptual framework of our study is based on the guidelines set out in the Nexus Working Group Brief: Policy issues in the financing of supply chains by Pete Baker et al. (3). They argue that health supply chains are a critical determinant of the performance of health systems and, by extension, of universal health coverage (UHC) accomplishment. They integrate two frameworks to understand the nexus between financing and logistics performance: UNICEF’s supply chain maturity model (Plan, Procure, Supply and Monitor) and WHO’s Public Financial Management (PFM) Cycle (budget formulation, execution, and monitoring)(3,4,5). The overlap between the two generates a matrix in which budget formulation is aligned with the planning phase, budget execution with procurement and delivery, and financial monitoring with logistical monitoring. This scheme situates the availability of inputs as the result of dynamic interactions between financial and operational decisions, an intersection addressed in this case study. Through this matrix, the study situates medicines availability as a direct outcome of dynamic interactions between financial and operational decisions.

This study analyzes the two financing models of the REMEDIAR Program from 2002-2024, focusing on budgetary planning, procurement efficiency, and supply chain performance. By examining four defined phases during this period, the analysis clarifies how distinct financing paradigms dictate different ’funding-procurement-distribution’ assembly strategies and their subsequent impact on program operation.

## Materials and Methods

### Design

This research employs a longitudinal single-case study (2002–2024), a methodology suited for addressing "how" and "why" questions within complex, real-world policy contexts (Yin, 6). By integrating quantitative and qualitative methods, the study analyzes the evolution of REMEDIAR’s financing, procurement, and distribution strategies.

### Data

The study employs a mixed-methods approach grounded in methodological triangulation, integrating primary and secondary sources to ensure a comprehensive analysis of the REMEDIAR Program (2002–2024). Qualitative evidence, derived from the thematic analysis of semi-structured interviews and normative documents, was used to reconstruct institutional milestones and identify the socio-political factors influencing program continuity. We also conducted a quantitative longitudinal analysis, using time-series methods to monitor financial and operational indicators over two decades. To ensure intertemporal comparability amid inflation and exchange rate volatility, budgetary data were re-expressed in US dollars and constant 2024 pesos using a spliced price index. Procurement efficiency was evaluated through the construction of the Awarded Price to Market Price (AP/MP) indicator, which served to estimate public savings and institutional bargaining power across different financing schemes.

Finally, findings were validated through data triangulation and expert feedback from key IDB officials to ensure that interpretations accurately reflected the program’s operational realities.

(see Table 1 Supplementary and Annex 1 for more information on interviews and REMEDIAR datasets).

### Phases under study

For the analysis, the study period was divided into four distinct phases based on the predominant funding source, even though these phases may overlap. This approach accounts for the different types of processes supported by the funding framework (FF). The specific phases chosen for this analysis are detailed in Figure 1.

**Figure 1.**
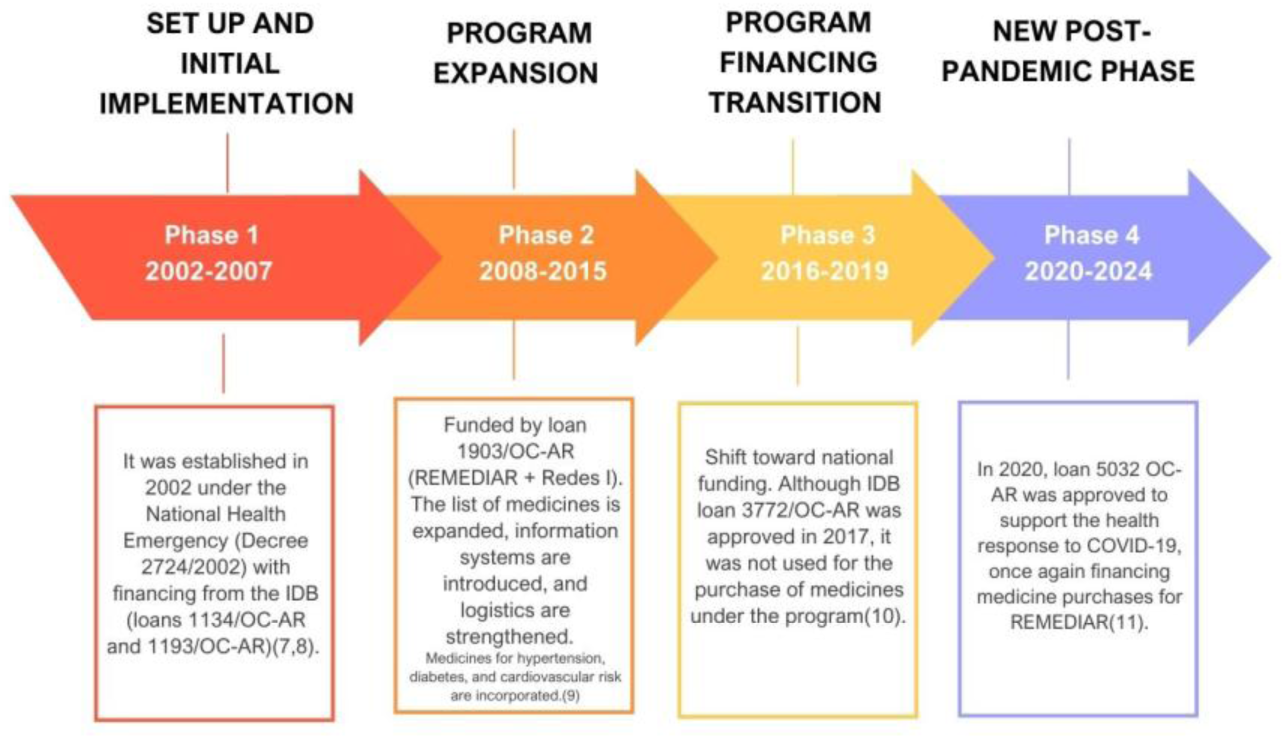
Phases for analysis.

### Ethics

The study did not require ethics committee review, as it was based on publicly available information, anonymized databases, and secondary sources. All interviews conducted included informed consent and confidentiality safeguards, in accordance with Law 25,326 on the Protection of Personal Data (12).

### Generative Artificial Intelligence (AI)

ChatGPT 4.0 was used to refine English style.

## Results

The REMEDIAR study seeks to operationalize the conceptual framework on health supply chain financing developed in the Nexus Working Group Brief (3).

## Funding

The REMEDIAR program has operated under two distinct financing models, each with its own institutional and operational frameworks, which have directly shaped its performance. Between 2002 and 2024, the budget followed a cyclical trajectory, closely tied to Argentina’s macroeconomic situation, fiscal space, and external credit availability.

Figure 2 illustrates the evolution of REMEDIAR’s budget in US dollars and its funding composition (2002–2024). The left panel reveals significant fiscal volatility, with peaks in 2008 and 2017. The right panel details the transition between funding mechanisms: Phase 1 (2002–2007) was characterized by modest national funding levels (green line) and a near-total reliance on IDB loans (pink line). This was followed by Phase 2, a period of sustained expansion driven by diversified funding and increased National Treasury participation, which facilitated broader program coverage. In contrast, Phase 3 saw a sharp contraction triggered by domestic fiscal constraints, currency depreciation, and austerity measures. This trend reversed abruptly in Phase 4 (post-2020); the pandemic’s onset prompted a significant budgetary rebound, sustained by a strategic combination of Treasury reinforcements and renewed access to international credit lines.

**Figure 2.**
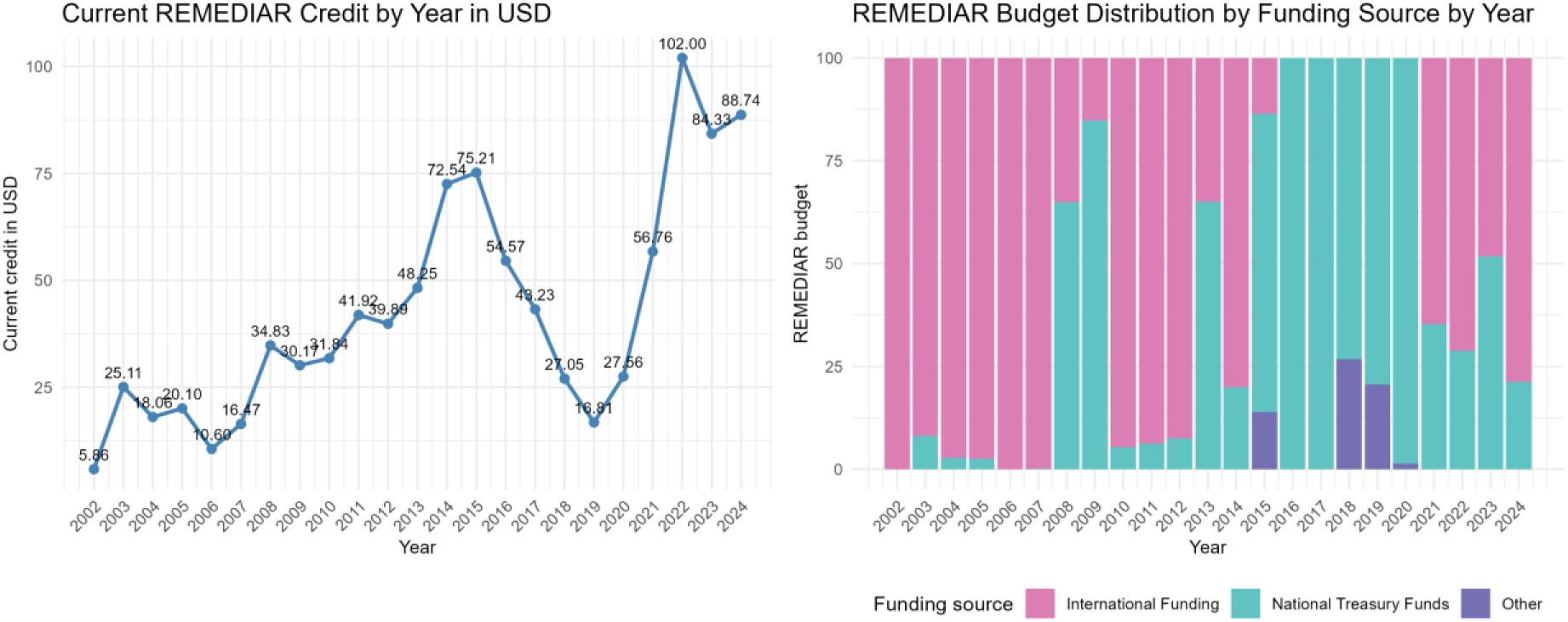
Budgetary Execution and Shifts in Resource Mobilization for the REMEDIAR Program (2002–2024). On the left: annual budgetary credit in US dollars. On the right: relative share of funding sources by fiscal year. The stacked bars represent the different financers: Inter-American Development Bank (IDB) international loans (pink), national treasury (green) and other sources (purple).

Budget execution represents the operational translation of allocated funds into actual spending. Between 2002 and 2024, REMEDIAR consistently achieved credit utilization levels of 90% or higher (Figure 3), demonstrating a robust capacity to transform allocations into deliverables. However, this performance was non-linear: following an initial start-up phase, efficiency surged in 2003–2004, with over-execution peaks in 2006 and 2021, and sharp declines in 2005, 2018, and 2022.

**Figure 3.**
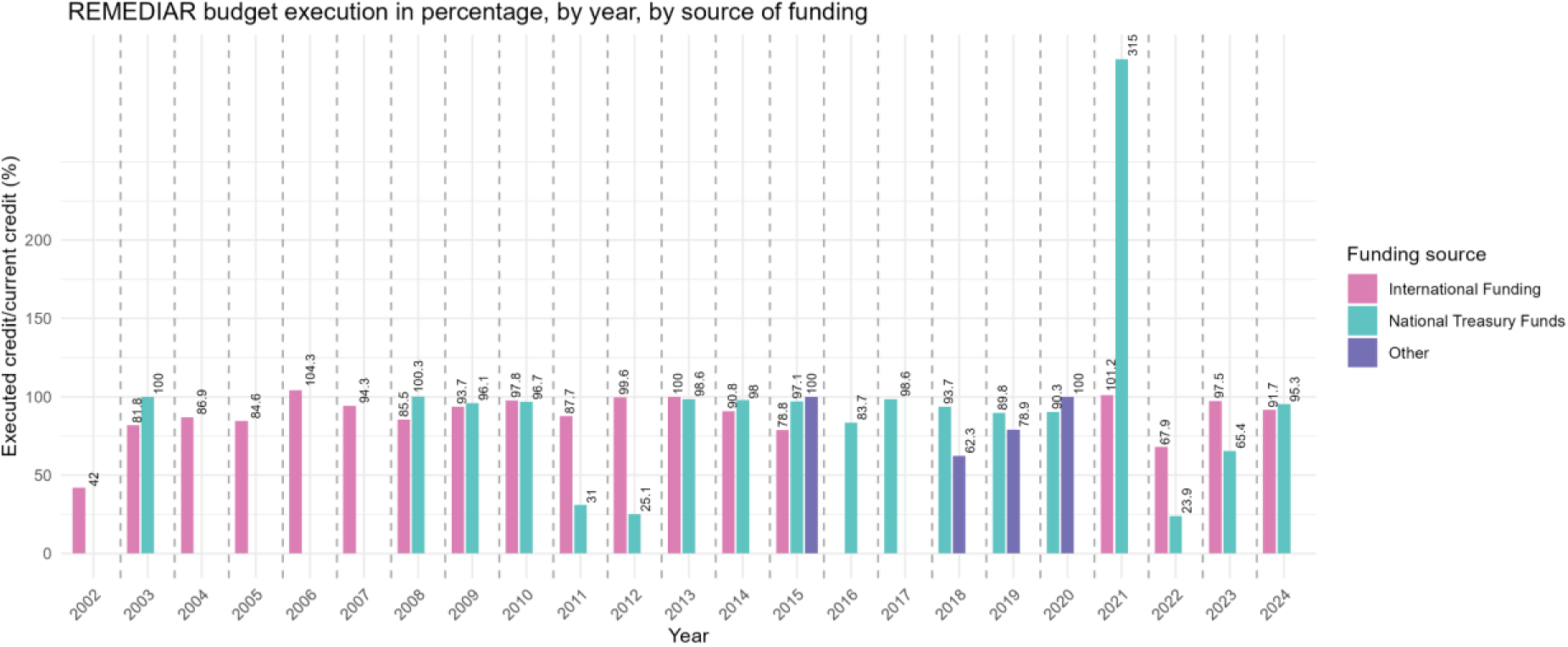
REMEDIAR budget execution in percentage, by year, by source of funding. The bars represent the different funding sources: Inter-American Development Bank (IDB) international loans (pink), national treasury (green) and other sources (purple).

Disaggregating these results by financing source reveals distinct operational dynamics. External credit from the IDB exhibits a consistent pattern of high execution, stabilizing above 80% and frequently exceeding 95% following the initial start-up period; such rates suggest that the IDB’s ring-fenced mechanisms effectively facilitate timely resource utilization. Conversely, the National Treasury’s performance is characterized by significant volatility; although it reached full execution in numerous periods, it suffered from marked under-execution in 2012 and 2022 due to domestic fiscal constraints. A notable outlier in this trend occurred in 2021, when execution exceeded 300% due to pandemic-related emergency reinforcements and extraordinary budgetary expansions. These divergent patterns highlight how funding sources shape the predictability and stability of program implementation over time.

While REMEDIAR has demonstrated political flexibility by remaining active across successive administrations, its budgetary history illustrates a persistent tension between social prioritization and Argentina’s macroeconomic constraints, making its institutional weight highly sensitive to sectoral priorities and the fiscal space.

### Procurement

Procurement timelines and time-series longitudinal data reveal systematic timing advantages under international financing across nearly all stages (Figure 4). National processes extended to a median of 458 days from initiation to purchase order (PO), whereas internationally financed processes did 48% faster, totaling 235 days. The most significant bottleneck in national schemes is the awarding–PO phase, which accounts for over half of the elapsed time (median: 249 days).

**Figure 4.**
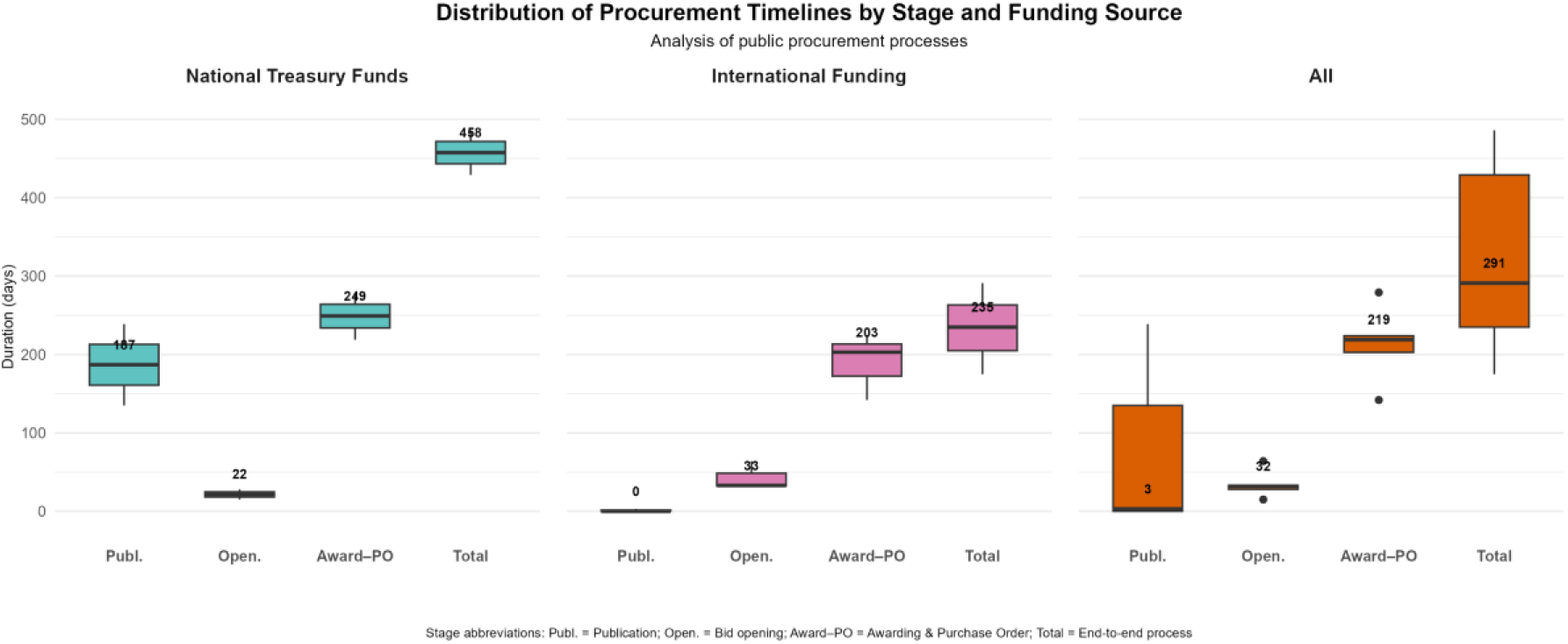
Distribution of procurement timelines by stage and funding source. Distribution of stage duration expressed in days for Publication, Bid Opening, Awarding, Purchase Order, and Total. Displayed separately for National and International financing, with an all panel pooling both schemes. Median values are labeled; boxes represent the interquartile range (IQR); whiskers extend to 1.5×IQR; points denote outliers.

The primary divergence occurs during the publication stage. In national processes, this phase is prolonged by sequential administrative approvals and dependence on immediate fund allocations. In contrast, international financing utilizes pre-approved credit lines, resulting in more stable timelines and a 40% reduction in the total publication-to-PO span. Although international bidding shows slightly longer opening times, these are offset by superior efficiency in subsequent stages.

The REMEDIAR strategy leverages centralized high-volume purchasing to secure significant savings relative to the retail market. To evaluate this, the Awarded Price to Market Price (AP/MP) ratio serves as a proxy for procurement efficiency, reflecting the discount the State achieves relative to the price paid by individuals.

Analysis of the 2002–2024 period reveals that the program began under exceptionally favorable conditions, with awarded prices at only 15% of market value in 2002, reaching a historical low of 9% in 2004. However, from 2008 (Phase 2) onwards, the AP/MP ratio shows an upward trajectory, signaling a gradual decline in relative efficiency. While international financing recorded peak ratios of 43%–50% (notably in 2006 and 2015), these spikes often reflected external currency fluctuations rather than diminished bargaining power. In contrast, national financing has maintained a more stable efficiency level between 21% and 30%, though it has not recovered the program’s early-year efficiency. These results underscore that while both models achieve substantial discounts, the international framework’s competitive conditions typically yield superior price optimizations for the most critical components of the essential medicine basket.

The Essential Medicines Formulary (VDM), the positive list of medications covered by REMEDIAR that address most of the health conditions treated at the PHC level, underwent significant programmatic expansion, growing from 39 active principles (51 presentations) in 2002 to 75 principles (99 presentations) by 2021. However, longitudinal data reveal a persistent gap between this theoretical basket and actual delivery (Figure 5).

**Figure 5.**
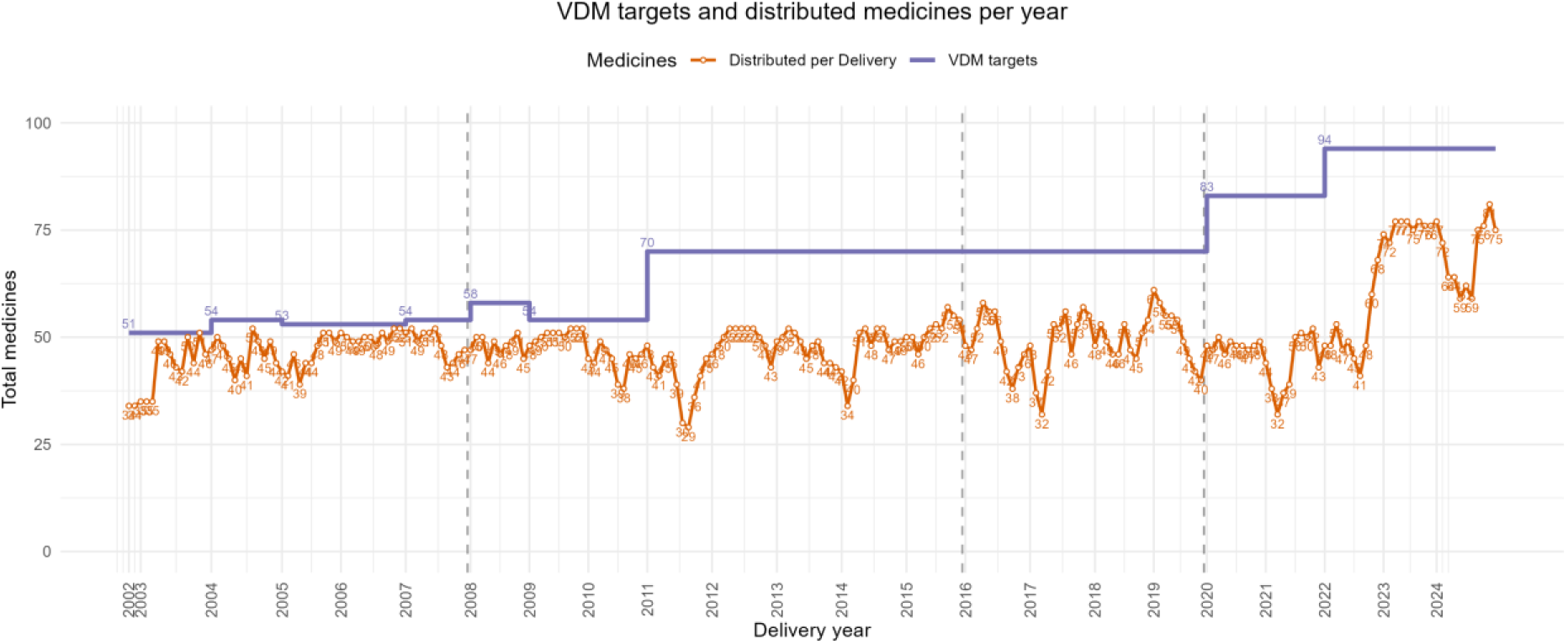
Number of types of medicines distributed per year (orange line) in comparison with the VDM targets as per ministerial resolutions (purple line). Along each year there are several distribution points, indicated in each orange point for the same year.

While the VDM targets followed a stepped growth—reflecting policy updates in 2010, 2019, and 2021 (REFS 13, 14, 15, 16)—the number of medicines effectively distributed (orange line) exhibits high variability. Although the VDM composition remained relatively stable, ensuring it was appropriate for primary care needs, the program struggled to ensure consistent fulfillment. Periods of near-total compliance were frequently followed by abrupt declines in the variety of distributed drugs.

These fluctuations were not driven by clinical changes but can arise due to various factors, such as budget constraints, failures in the bidding processes, limited market availability, or significant discrepancies between reference prices and submitted bids.

The resulting gap highlights that the VDM, while technically robust, often exceeds the program’s operational capacity to sustain a complete and uninterrupted supply chain.

In theory, decentralized procurement models offer greater agility and flexibility to adapt to local epidemiological changes; however, they often entail the erosion of economies of scale and significant price dispersion across purchasing entities. This study illustrates this tension along two dimensions: the incorporation of medicines into the Essential Medicines Formulary (VDM) and their effective procurement for specific pathologies proved more complex than in other models based on decentralized purchasing decisions. A key example is the integration of mental health medicines: although the strategic decision was adopted in 2018, full implementation—including inclusion in the formulary and the execution of pilot trials—required four years. This highlights a critical trade-off: while centralization ensures fiscal sustainability (as discussed in the following section), it may delay the alignment of supply with emerging clinical demands.

To further analyze procurement efficiency under the REMEDIAR Program, a comparative analysis was conducted using a set of essential high-volume medicines. Empirical data confirms that centralized procurement acts as a powerful rationalizing tool. For a comparable basket of essential medicines, REMEDIAR’s centralized acquisitions averaged 25% of market prices, whereas decentralized purchases at the subnational level reached 46%—nearly double the cost, using the district of the city of Buenos Aires as a more reliable data proxy for subnational purchasing. (Table 1).

**Table 1.** AP/MP ratio (% of market price) for a common basket of medicines: decentralized (health centers) vs. centralized (REMEDIAR). Values are expressed as % of market price. Simple averages by medicine. Decentralized figures correspond to public-tender purchases; gaps are likely conservative relative to non-tender modalities.

| Drug | Average (AP/MP) |  |
| --- | --- | --- |
|  | Health center | REMEDIAR |
| **Azithromycin** | **50%** | **21%** |
| **Carbamazepine** | **72%** | **16%** |
| **Fluconazole** | **49%** | **58%** |
| **Levodopa + Carbidopa** | **95%** | **31%** |
| **Levothyroxine** | **87%** | **25%** |
| **Paracetamol** | **28%** | **19%** |
| **Benzathine Penicillin G** | **30%** | **16%** |
| **Sodium Divalproate** | **79%** | **19%** |

A variation in unit prices, within the same jurisdiction and for the same product, underscores structural limitations of decentralized models in achieving price efficiency. Overall, the pattern reinforces the redistributive and rationalizing role of centralized purchasing, not only as a tool for fiscal optimization, but also as a mechanism for reducing access disparities and strengthening institutional equity.

### Distribution

The Nexus conceptual framework notes that last-mile delivery is the most neglected phase from a financing perspective, despite its very high costs (storage, transportation, logistics). In this regard, the REMEDIAR study provides empirical evidence on the outsourcing of logistics—storage, kit assembly, and transportation—to specialized private operators, combined with centralized distribution and direct delivery to healthcare facilities. It demonstrates that, although the logistics system achieved high standards (ISO certification, traceability), its sustainability is limited by administrative complexity and a lack of flexibility to adapt deliveries to emergencies or emerging epidemiological needs.

The implementation of logistics within REMEDIAR entails complexity due to its territorial scope in a very large country like Argentina (3.669.711 km), the distribution to more than 8.000 PHC facilities across all 24 jurisdictions, the use of medicine kits tailored to the specific needs of each territory (with more than 400 different kit configurations deployed), and the multiplicity of actors involved. Two major policy decisions were adopted in the design phase of the program. First, the direct distribution of medicine kits from the central level to each PHC, without the intermediation of jurisdictional warehouses, thereby avoiding delays, operational difficulties in local redistribution, and potential discretionary uses.

The second decision concerned the outsourcing of logistics services for medicine storage, kit assembly, and distribution. According to the logistical cost of the REMEDIAR program (18), the reasons for subcontracting logistics included cost reduction, improved service levels, and access to external infrastructure and technology required to ensure product storage and regulatory compliance. Nonetheless, although logistics operations are outsourced, the REMEDIAR Program remains accountable for logistics as a whole, being responsible for demand planning, quality supervision, information and traceability systems management, and coordination with jurisdictions, while the private sector is in charge of specialized storage, inventory management, differentiated kit assembly, and the physical distribution of medicines.

Figure 6 shows the distribution of treatments by province for 2023. The first map, which presents the total distribution of treatments in absolute terms, the province of Buenos Aires stands out clearly, with more than six million treatments. The second map relates treatments to the total population, revealing a higher distribution concentration in provinces such as Formosa, Santiago del Estero, Chaco, and Catamarca, all reporting values exceeding 1.5 treatments per inhabitant. The third map, which adjusts distribution based on the population with Exclusive Public Coverage (i.e., individuals without social security or private health insurance), shows that northern and northeastern provinces have the highest levels (exceeding three treatments per person with public coverage), showing that a significant proportion of treatments is concentrated in the regions that have historically faced more vulnerable health conditions.

**Figure 6.**
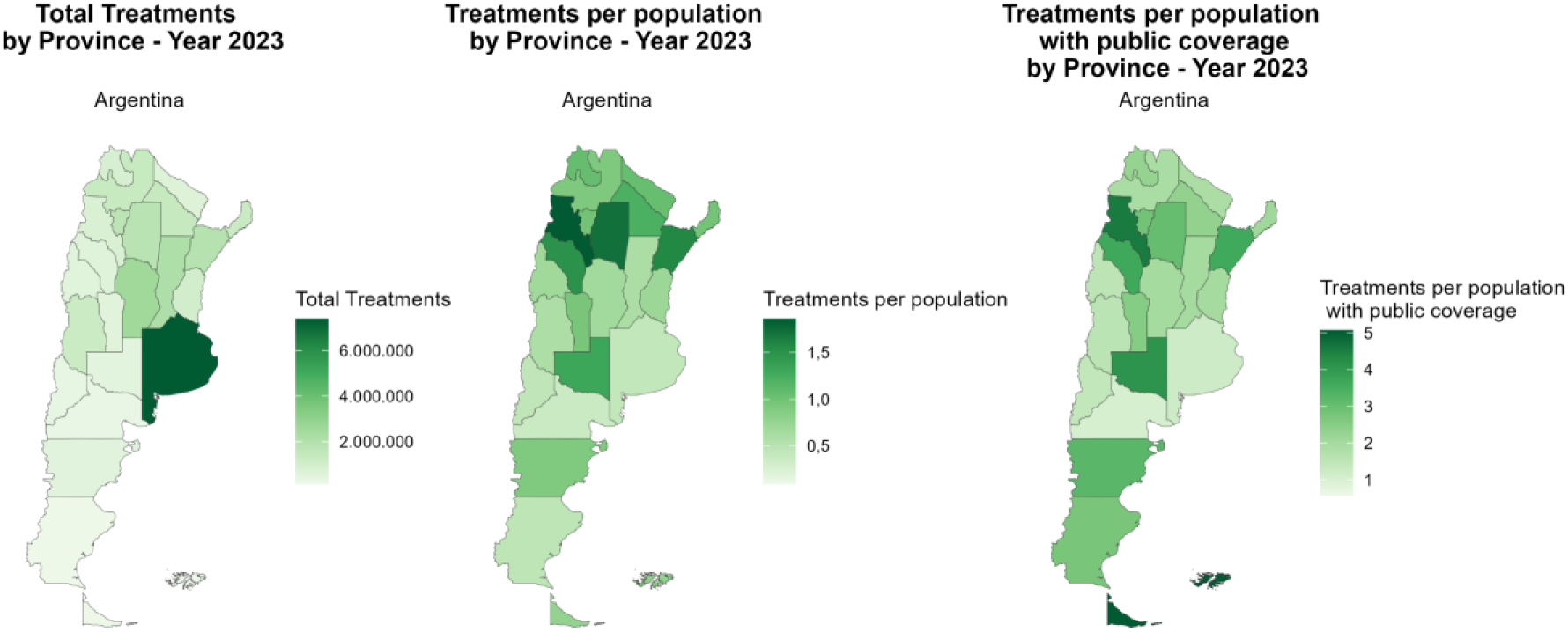
Distribution of treatments by province, year 2023. On the left: total treatments. In the middle: treatments per province population. On the right: treatments per population with public health coverage.

Beyond improving geographical equity in access, the scale and continuity of medicine distribution under REMEDIAR have direct implications for household financial protection, particularly for uninsured populations.

### Out-of-Pocket spending and Financial protection

Out-of-pocket (OOP) health expenditures are a key indicator of financial protection in health systems, as they represent direct payments households must make at the point of care. According to the World Health Organization (WHO), this type of spending can lead to access barriers and impoverishment, especially among low-income populations (19).

In Argentina, medicines account for the largest share of out-of-pocket health expenditures. By 2017, they accounted for approximately 38% of the household’s health spending.

Figure 7 presents medicine spending across income quintiles in Argentina at three points in time: before the Program (1997), during the early implementation phase (2004), and in the most recent household spending survey (2017-2018).

**Figure 7.**
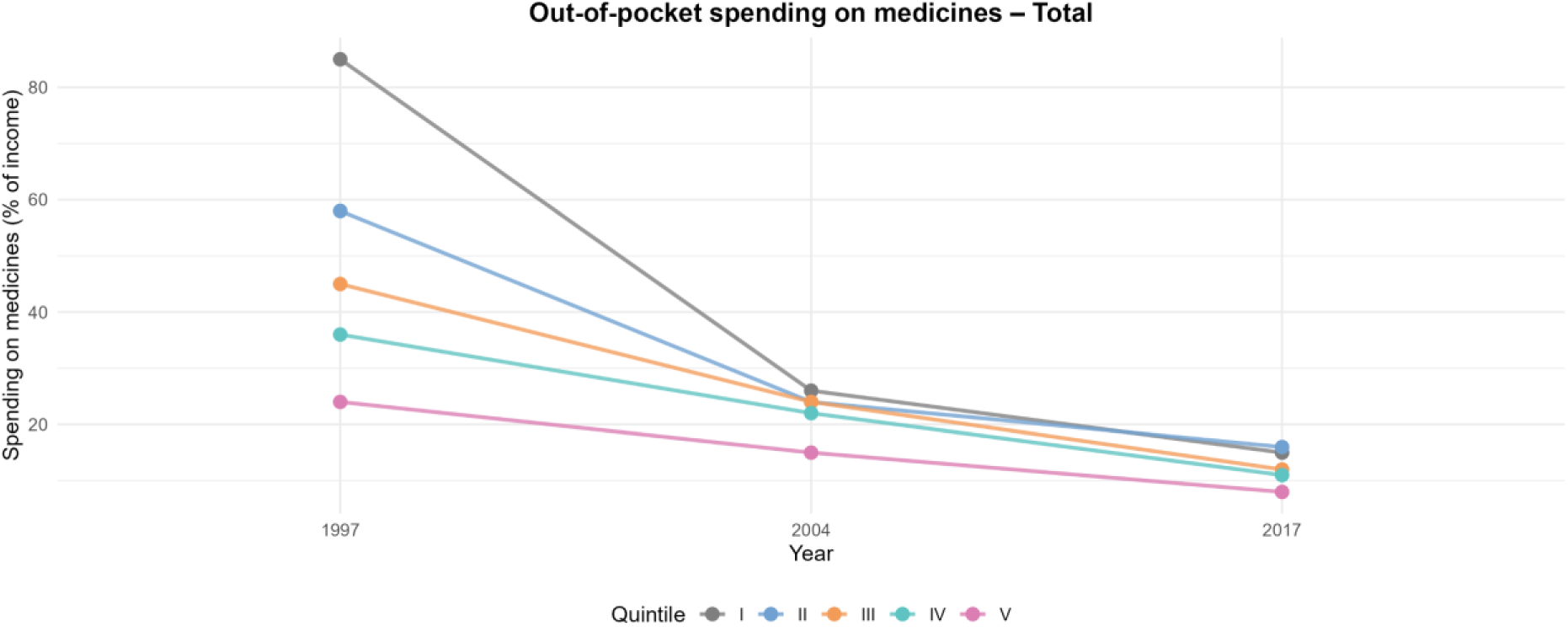
Spending on medicines by quintiles in Argentina.

Analysis of the initial indicators reveals a regressive pattern in out-of-pocket spending on medicines: lower-income households allocate a significantly higher proportion of their income to the purchase of medications. The implementation of the REMEDIAR Program over time is associated with a substantial reduction in this financial burden, with particularly visible effects in the lower income quintiles.

Between 1997 and 2004, the financial effort required to obtain medicines decreased markedly across all income quintiles. In the first quintile, the burden fell from around 9% of household income in 1997 to approximately 2–3% in 2004, representing the largest absolute reduction in the period. Middle- and upper-income quintiles also experienced consistent declines, converging toward levels below 2%. By 2017, out-of-pocket spending remained relatively low, stabilizing around 1.0–1.6% of household income across quintiles.

Taken together, these results show a generalized downward shift in the financial effort associated with the purchase of medicines. However, the magnitude of the reduction varies across income groups, reflecting differences in the sensitivity of household spending to the increased availability of free treatments provided by the REMEDIAR Program and to the economic conditions affecting each income stratum.

The evolution of out-of-pocket spending on medicines across the periods 2004–2005, 2012–2013, and 2017–2018 shows substantial variations in the economic burden borne by households covered by the Program (Figure 8).

**Figure 8.**
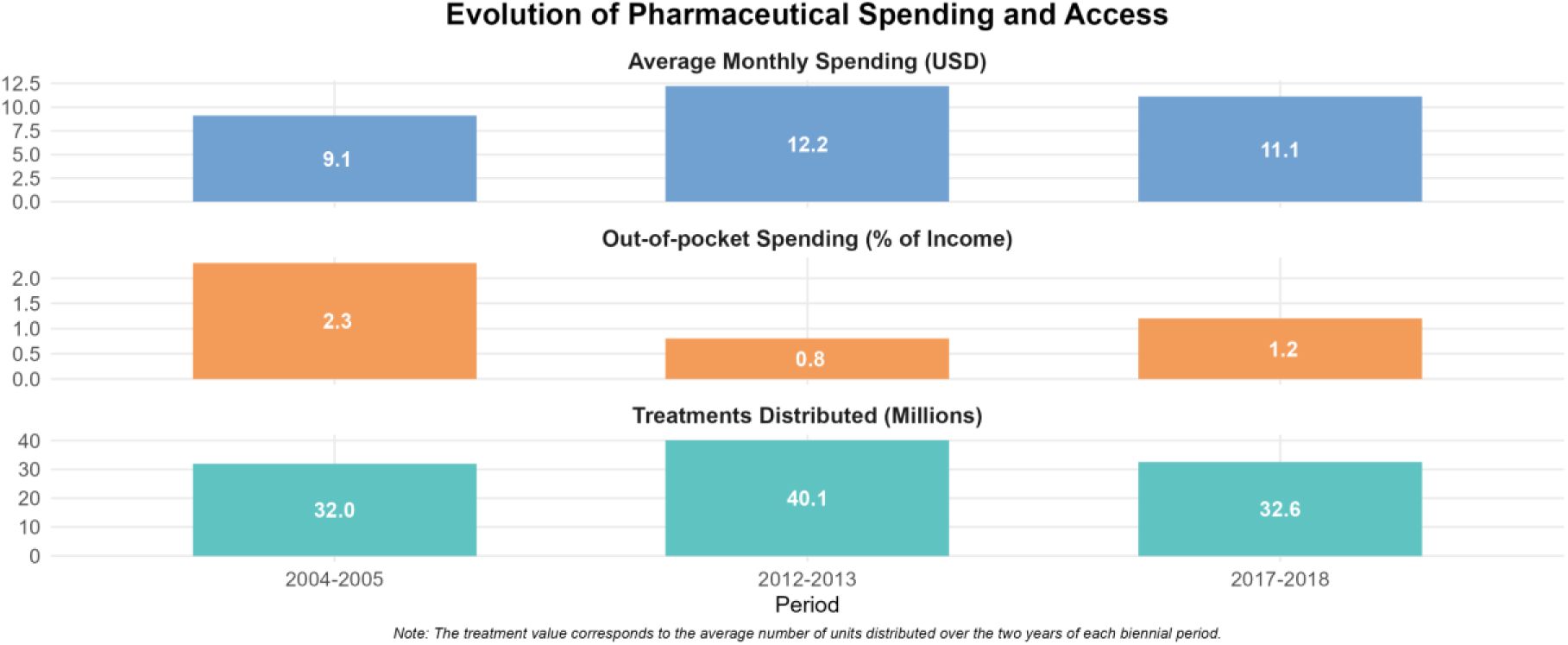
Trends in Spending, Treatments, and Out-of-Pocket Medicine Costs

Figure 8 reports average monthly household OOP spending on medicines (USD) and its share of household income for the three periods, displayed alongside the annual number of treatments distributed by REMEDIAR. Labels indicate reference values for each period The empirical evidence suggests an inverse relationship between program intensity—measured by the volume of treatments distributed—and the financial burden of medicine spending. The greater the availability of free-of-charge treatments, the lower the economic strain on uninsured households. From a public policy perspective, these findings highlight the countercyclical nature of the REMEDIAR Program, as it helps to cushion the effects of declining incomes during regressive periods. At the same time, they underscore the vulnerability of this protection in the face of fiscal tightening or operational constraints.

## Discussion

The two-decade trajectory of the REMEDIAR Program provides a unique opportunity to analyze the intersection of public financial management (PFM) and health supply chain performance. By applying the Nexus conceptual framework, this study demonstrates that the availability of essential medicines is not merely a logistical outcome but a direct product of the financial architecture underpinning the system.

The analysis of REMEDIAR’s funding cycles reveals a persistent "financing duality". International concessional loans (IDB) acted as a stabilizing force, providing a "protective envelope" that shielded the program from Argentina’s recurring macroeconomic crises. This stability is rooted in what we define as the international financing package: payment in hard currency, earmarked funds, and performance-contingent disbursements.

These conditions effectively reduced suppliers’ risks. When contracts are denominated in US dollars and payments follow predictable timelines, the private sector lowers its risk premiums, leading to the significantly lower prices observed in Phases 1 and 4. In contrast, reliance on the National Treasury introduced "fiscal noise." Despite the Treasury’s high execution rates in stable years, its exposure to currency devaluation and to discretionary austerity measures in Phases 2 and 3 led to "budgetary fragmentation." This fragmentation forced the program to reconcile disparate rules and timelines, ultimately compromising the continuity of supply. The findings suggest that while domestic funding is politically sovereign, it lacks the built-in discipline of external credit in LMIC, creating a dependency trap where international loans become the only viable path to efficiency.

In relation to procurement, REMEDIAR provides compelling evidence for the redistributive role of centralized procurement. In a federal country with vast asymmetries in fiscal capacity like Argentina, centralization is a tool for equity, though it requires a strong national state capacity. By consolidating demand, the National Ministry of Health offsets the administrative weaknesses of smaller, poorer provinces, ensuring that a citizen in a remote region has access to the same essential basket of goods as one in a wealthy urban center. However, our empirical comparison between centralized and decentralized models reveals a profound trade-off. Centralization achieves an average price efficiency of 25% relative to market value—nearly double the efficiency of decentralized hospital purchases (46%). Yet, this fiscal optimization comes at the cost of institutional rigidity. The four-year lag in integrating mental health medications into the VDM illustrates how centralized administrative "gravity" can slow down clinical adaptation. The Nexus framework’s tension between scale and agility is manifest here: the same bureaucratic density that secures the lowest price also acts as a barrier to innovation. For the program to evolve, it must find a "hybrid governance" model that maintains centralized bargaining power while decentralizing the feedback loop for epidemiological updates.

A key contribution of this study is the evidence regarding the outsourcing of the "last mile." REMEDIAR’s decision to bypass jurisdictional warehouses for direct delivery to over 8,000 Primary Health Care (PHC) centers was a strategic move to prevent local discretionary use and redistribution delays. This "direct-to-clinic" model, supported by ISO-certified private operators, represents the highest standard of pharmaceutical traceability in the region. Nonetheless, the study identifies a structural vulnerability in the outsourcing model. The competitive bidding process for logistics services, while ensuring cost-efficiency, creates "transition shocks." Every time a contract changes hands, the entire operation—warehousing, inventory, and kits—must be relocated. These transitions have historically caused supply interruptions, highlighting that the program is "accountable but not operational." The reliance on private infrastructure, while technologically superior, leaves the State with limited contingency options if a provider fails or a contract expires. The sustainability of the "last mile" thus depends on a more sophisticated regulatory capacity that can manage these transitions without compromising patient access.

The analysis of out-of-pocket spending confirms that REMEDIAR’s contribution to equity extends beyond access, operating as a countercyclical financial protection mechanism. Periods of budgetary expansion and high distribution volumes coincide with the lowest levels of household financial burden, while fiscal contraction translates into partial erosion of this protection.

The transition from international to national financing (the "phasing out" process) has historically been treated as a budgetary task rather than an institutional redesign.

Based on twenty years of evidence, we propose three key policy shifts:

1. Internalization of IDB Best Practices: protecting drug resources and multi-year budget to provide predictability over time.
2. Formal Federal Governance: the federal health council should serve as a governance structure allowing REMEDIAR to move from a centralized command-and-control model where provinces should be formally integrated into the decision-making process—not just as recipients of kits, but as partners in VDM updates and demand planning. This would align national supply with local epidemiological realities and reduce the 4-year lag in integrating new therapies.
3. Demand-Driven Logistics: The program must transition from a "supply-push" model (delivering what is available) to a "demand-pull" system (delivering what is consumed). This requires investing in real-time digital stock management at the PHC level to eliminate the "hidden demand" problem where physicians stop prescribing due to perceived stockouts.

### Limitations

The findings of this study are subject to methodological limitations, including the heterogeneous quality of early administrative records (2002–2005), potential recall bias in retrospective interviews, and uneven provincial data availability for comparison between centralized and decentralized purchasing, except for the district of the city of Buenos Aires, which was considered a more reliable subnational proxy in terms of data quality and completeness.

## Conclusion

The REMEDIAR program fundamentally transformed the procurement and distribution of essential medicines in Argentina, shifting from a highly fragmented and decentralized model to a centralized system. Prior to its implementation, provinces operated independently, resulting in marked inequalities in access between wealthier and poorer regions, with the latter facing chronic shortages. The introduction of REMEDIAR established centralized purchasing, direct distribution to primary health centers, and a comprehensive audit system, which together significantly improved the national supply of medicines.

The analysis presented in this paper confirms that international financing—primarily through loans from the Inter-American Development Bank (IDB)—was more efficient than national financing during the period 2002–2024. This was evident in several key areas, including procurement timelines, predictability, awarded prices, and budget execution. The analysis also raises possible explanations for this relative efficiency. In this context, the program’s long-term sustainability is challenged, particularly during the transition from international to national financing. Success depends on aligning health priorities with government budget allocations, a process that requires political decisions and trade-offs. Sustainability goes beyond simply replacing funds; it entails strengthening governance, financing, and service delivery across the entire health system.

In Argentina, the transition towards national financing was closely tied to broader structural political changes that imposed significant budgetary constraints. The consolidation of nationally financed structural policies undoubtedly requires well-established practices that prioritize administrative efficiency in procedures, forward-looking planning, and the strengthening of appropriate monitoring and oversight mechanisms. These requirements are not inherently linked to international processes but rather to sustainable practices and the availability of adequate domestic resources. Otherwise, equating efficiency solely with international financing risks distorting the analysis and may lead to premature conceptualizations.

The analysis of the REMEDIAR Program offers lessons that go beyond the Argentine case and are relevant for other countries in the region facing similar tensions between international and national financing. Experience shows that the efficiency observed under international credit schemes does not depend exclusively on the availability of external resources, but rather on the associated management rules.

## Annex 1

### Re-expression of values as a comparability criterion

Budget credits and uses are recorded in current pesos. To enable intertemporal comparisons and isolate nominal effects, the series are re-expressed along two complementary analytical domains: current U.S. dollars, as an international unit of account. and constant 2024 pesos as the real measure. The USD view is used strictly as a magnitude and external-exposure reference; real variations are assessed in ARS 2024. The reference exchange rate used was from BCRA Communication ‘A’ 3500.

### Deflation to constant prices

Real measurement is performed in 2024 pesos, using the Consumer Price Index (general level) as the deflator. Owing to well-documented methodological inconsistencies in official statistics between 2007 and 2015, a spliced CPI series is constructed and normalized so that 2024 = 100.

Splicing strategy (annual Dec/Dec rates):

- 2001–2006: annual Dec/Dec variations from CPI-GBA (Greater Buenos Aires), the only official reference available for that period.
- 2007–2016: simple average of the Dec/Dec annual variation rates published by CPI-CABA, Mendoza, and San Luis, selected for their technical soundness, continuity, and institutional independence.
- 2017–2024: Dec/Dec annual variations from the National CPI (INDEC), once methodological reliability was re-established.

For the 2007–2016 splice, we use the simple average of Dec/Dec annual rates from CPI-CABA, Mendoza, and San Luis, selected for their robustness, reliability, and data availability. Regional indices do not exhaust national coverage, but their combination yields a stable and methodologically consistent approximation for a centrally executed program with relatively homogeneous price baskets.

### Efficiency in procurement processes

1. Unit of analysis: molecule–dose expressed in the Unit of Minimum Dispensing (UMD)
2. Market price source and event dating The market price (MP) is drawn from the Alfabeta Pharmaceutical Manual, a reference that records retail pharmacy prices (PVP) prevailing in the domestic market.
3. Study window and inclusion/exclusion criteria The analytical window is 2002–2018, selected for data availability/consistency and to ensure a stable institutional environment for structural assessment. Subsequent years are excluded due to the COVID-19 shock, which profoundly altered relative prices, procurement modalities, and introduced administrative discontinuities that break time-series comparability. To maintain methodological homogeneity and focus on competitive procedures, direct contracting is excluded; the evaluation concentrates on public tenders with awards.
4. Decentralized benchmark: CABA (2015–2018)

**Table 1 SUPL:**
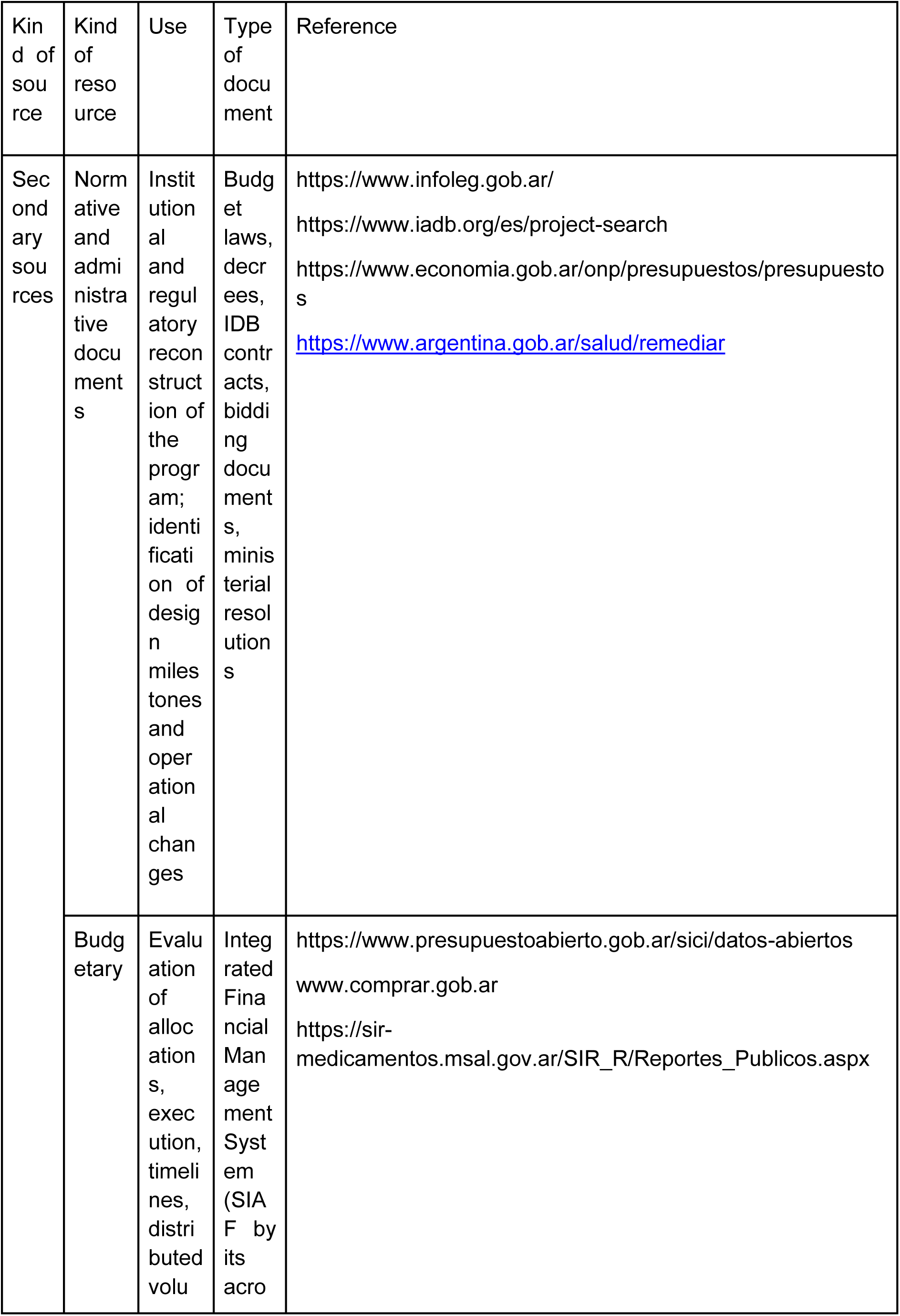
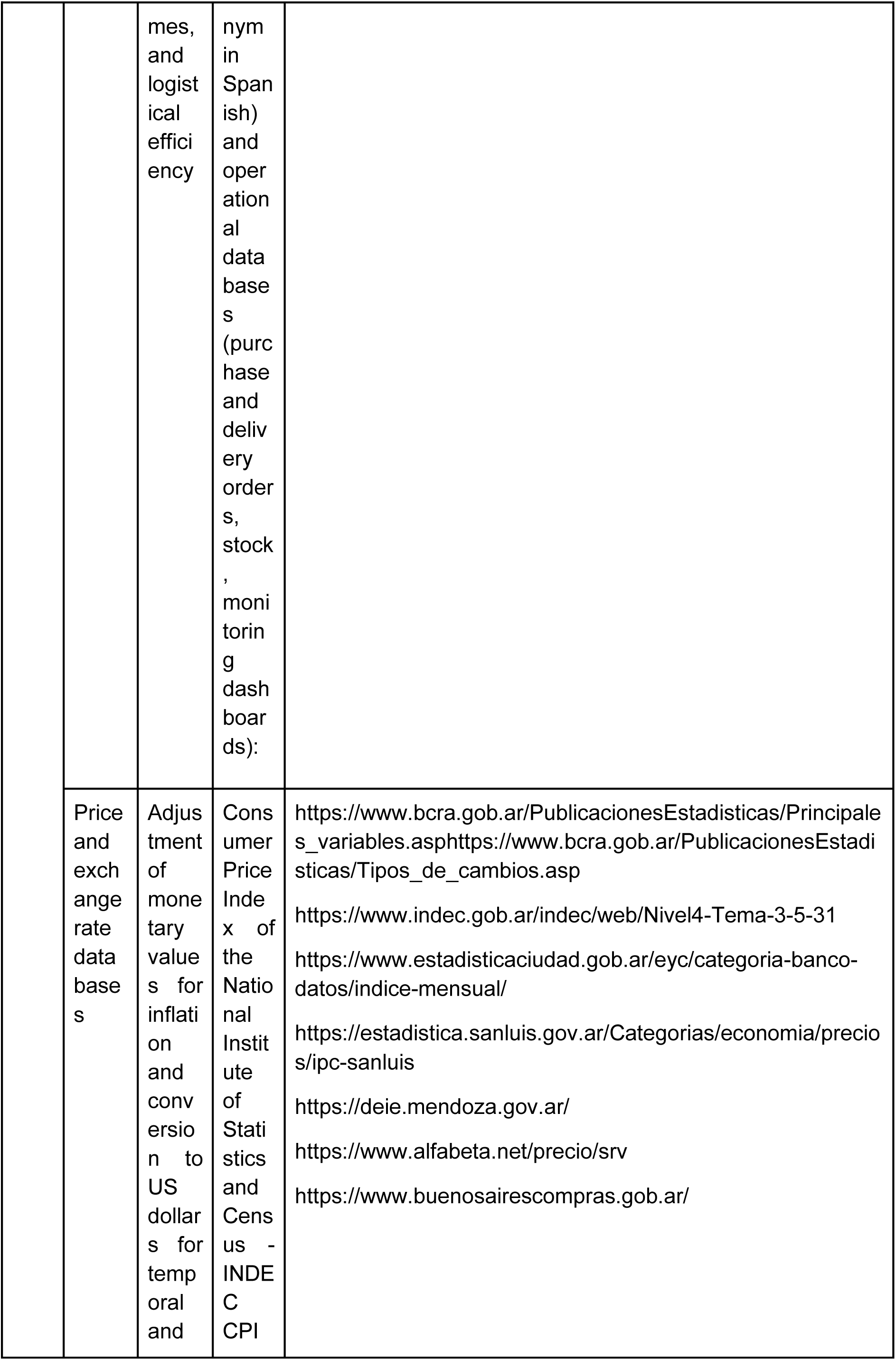
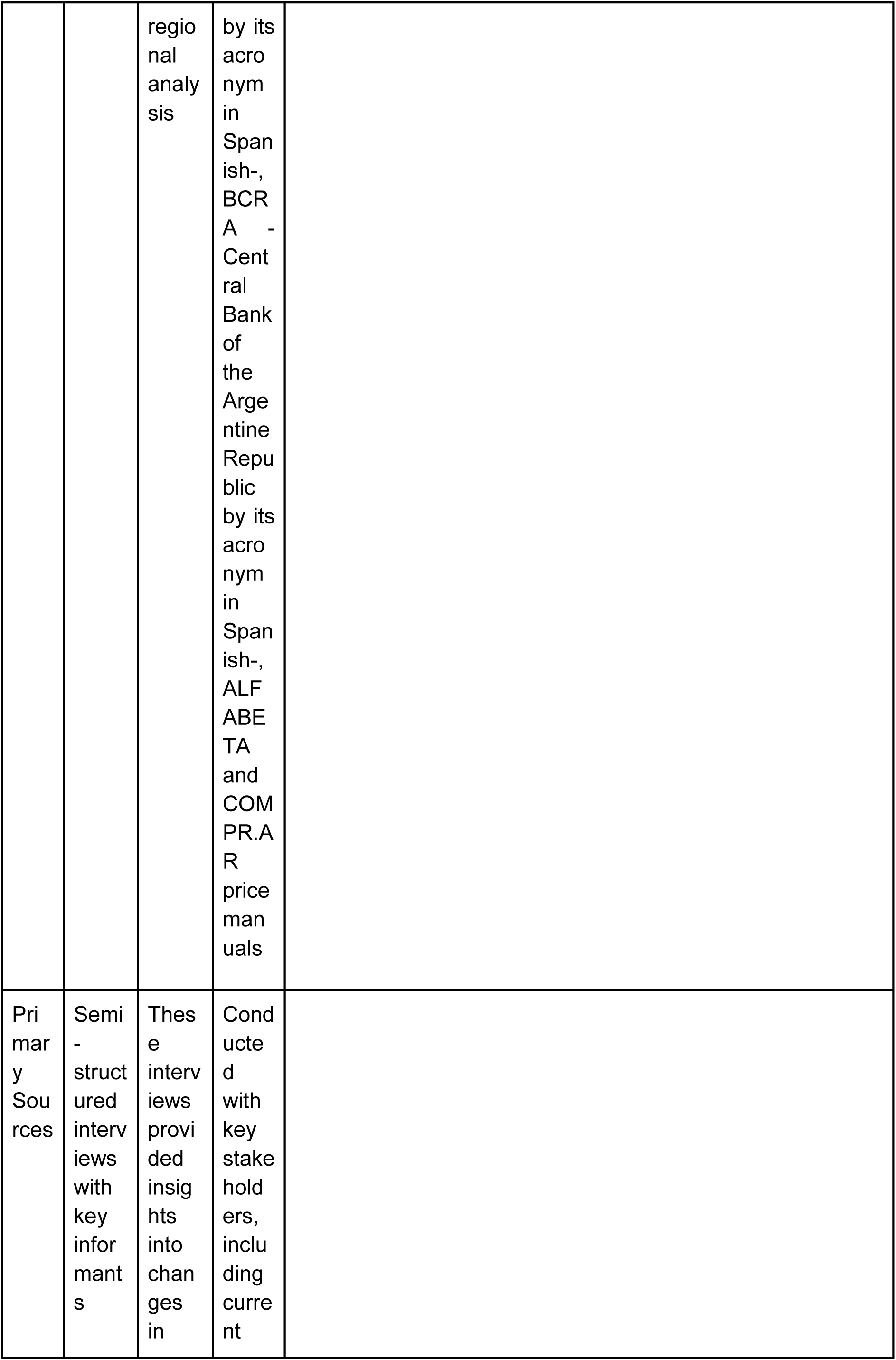
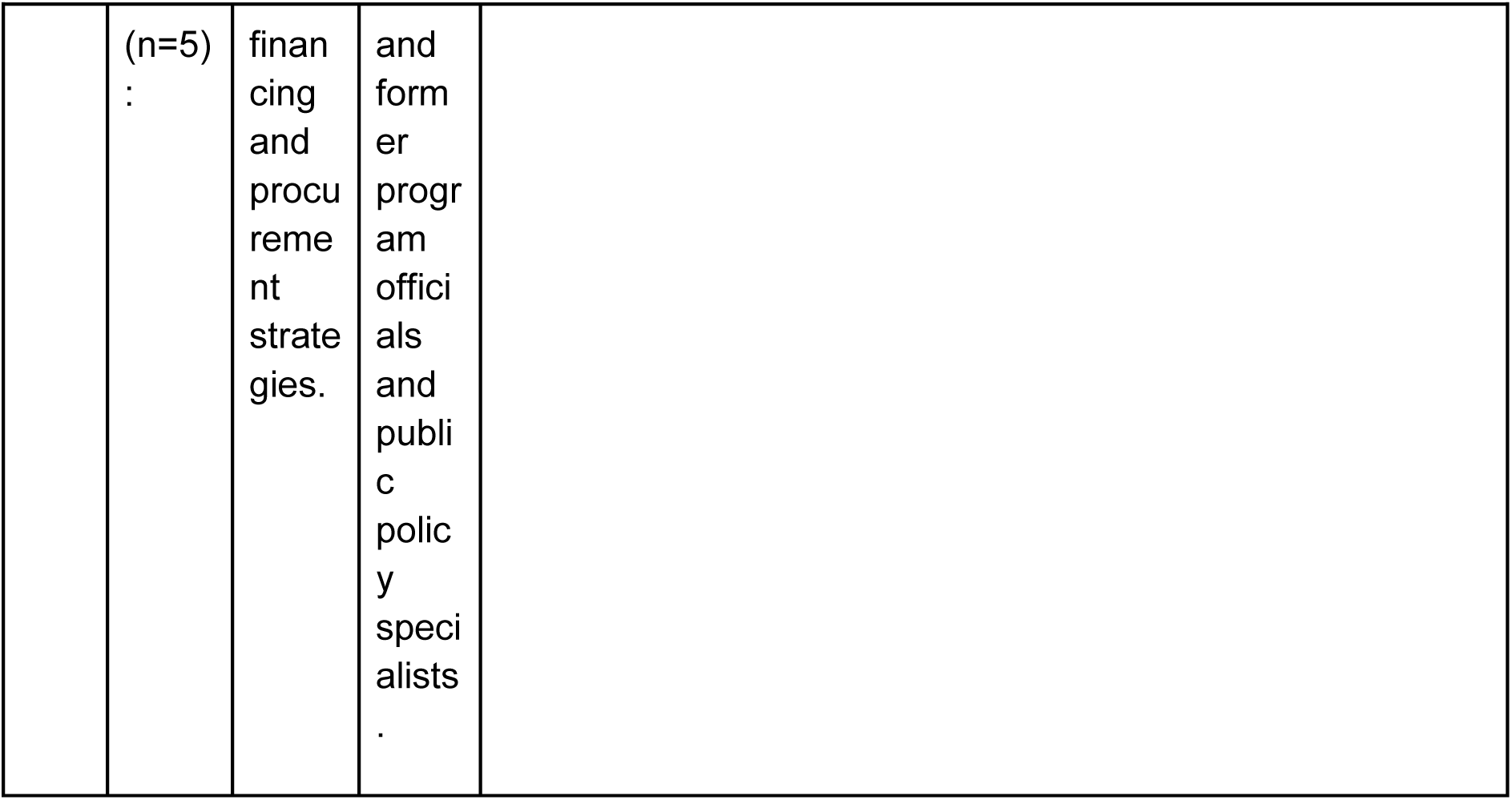
Sources of information used for methodological triangulation.

## Data availability statement

The data that support the findings of this study are available in essentialMedicines at https://github.com/LuciaBart/essentialMedicines. These data were derived from the following resources available in the public domain: https://www.presupuestoabierto.gob.ar/sici/datos-abiertos<u>#</u>, https://www.indec.gob.ar/indec/web/Nivel4-Tema-4-45-151, https://www.alfabeta.net/precio/srv, https://comprar.gob.ar/Default.aspx, https://comprar.gob.ar/Default.aspx, https://www.argentina.gob.ar/salud/remediar/medicamentos-esenciales, https://www.buenosairescompras.gob.ar/, https://sir-medicamentos.msal.gov.ar/SIR_R/Reportes_Publicos.aspx

